# Pancreas MRI segmentation into head, body, and tail enables regional quantitative analysis of heterogeneous disease

**DOI:** 10.1101/2021.11.30.21266158

**Authors:** Alexandre Triay Bagur, Paul Aljabar, Gerard R Ridgway, Michael Brady, Daniel P Bulte

**Affiliations:** Department of Engineering Science, University of Oxford, Oxford, UK; Perspectum Ltd, Oxford, UK

**Keywords:** Segmentation, groupwise registration, NAFPD, diabetes, MRI-PDFF, heterogeneity

## Abstract

Pancreatic disease can be spatially inhomogeneous. For this reason, quantitative imaging studies of the pancreas have often targeted the 3 main anatomical pancreatic segments, head, body, and tail, traditionally using a balanced region of interest (ROI) strategy. Existing automated analysis methods have implemented whole-organ segmentation, which provides an overall quantification, but fails to address spatial heterogeneity in disease. A method to automatically refine a whole-organ segmentation of the pancreas into head, body, and tail subregions is presented for abdominal magnetic resonance imaging (MRI). The subsegmentation method is based on diffeomorphic registration to a group average template image, where the parts are manually annotated. For a new whole-pancreas segmentation, the aligned template’s part labels are automatically propagated to the segmentation of interest. The method is validated retrospectively on the UK Biobank imaging substudy (scanned using a 2-point Dixon protocol at 1.5 tesla), using a nominally healthy cohort of 100 subjects for template creation, and 50 independent subjects for validation. Pancreas head, body, and tail were annotated by multiple experts on the validation cohort, which served as the benchmark for the automated method’s performance. Good intra-rater (Dice overlap mean, Head: 0.982, Body: 0.940, Tail: 0.961, N=30) as well as inter-rater (Dice overlap mean, Head: 0.968, Body: 0.905, Tail: 0.943, N=150) agreement was observed. No significant difference (Wilcoxon rank sum test, DSC, Head: p=0.4358, Body: p=0.0992, Tail: p=0.1080) was observed between the manual annotations and the automated method’s predictions. Results on regional pancreatic fat assessment are also presented, by intersecting the 3-D parts segmentation with one 2-D multi-echo gradient-echo slice, available from the same scanning session, that was used to compute MRI proton density fat fraction (MRI-PDFF). Initial application of the method on a type 2 diabetes cohort showed the utility of the method for assessing pancreatic disease heterogeneity.

## 1. Introduction

### 1.1. Heterogeneity of chronic pancreas disease

Pancreas pathology, such as fatty infiltration, diabetes, chronic pancreatitis, and pancreatic cancer, are rising rapidly with the increasing prevalence of obesity and the metabolic syndrome. Obesity leads to ectopic fat deposition in organs like the heart, liver, and pancreas. While non-alcoholic fatty liver disease (NAFLD) is a well-recognized disease entity, now affecting 1/4^th^ of the worldwide population and 1/3^rd^ of US adults [1], non-alcoholic fatty pancreas disease (NAFPD) was only coined relatively recently [2], [3] despite showing similar prevalence in a meta-analysis [4]. Analogously to NAFLD, NAFPD triggers inflammatory processes that, if left untreated, may lead to chronic pancreatitis and pancreatic cancer [5], [6]. NAFPD has also been linked to type 2 diabetes [7], [8]. Early detection of pancreatic disease is therefore important, however these are often “silent” conditions that only become symptomatic at a late stage, when they may already be untreatable. Incidental findings, where the target organ is near the pancreas, for instance in quantitative imaging of the liver, potentially offer a way to detect pancreas pathology in time.

Pancreatic disease processes, including fat infiltration, fibro-inflammation, and pancreatic cancer, are also spatially inhomogeneous [9], [10]. There is increasing interest in studying pancreatic disease and the implications of disease heterogeneity, aiming to describe regional differences and localize pancreatic lesions. Early work using computed tomography (CT) classified uneven pancreatic fat infiltration into multiple subtypes or patterns, depending on the affected regions [10]. Uneven distribution of islet cells, that are responsible for insulin secretion and blood sugar regulation, has been reported using histology [11]. Fibrosis was more commonly found in the ventral pancreas than in the dorsal pancreas in patients with ampullary carcinoma [12]. The frequency of pancreatic cancer also differs regionally, with 60-70% occurrence in the head of the pancreas, and the symptoms vary by the location [13], [14]. From the imaging modalities commonly used for pancreatic assessment, including histology, endoscopic ultrasound, contrast-enhanced CT, and magnetic resonance imaging (MRI), only MRI can provide non-ionizing, non-invasive quantitative information of pancreas state, while providing full coverage and measures of spatial heterogeneity. Quantitative MRI biomarkers such as proton density fat fraction (PDFF) and T1 have shown potential in detecting pancreas steatosis and early-stage chronic pancreatitis, respectively [15], [16]; PDFF has been used for longitudinally monitoring total pancreatic fat deposition in a diabetes remission trial [17].

Apparent diffusion coefficient (ADC) from diffusion-weighted imaging has shown potential at grading malignancy of a certain pancreatic neoplasm type [18]. While some studies using MRI reported clinically important quantitative differences between pancreas subsegments [19], [20], other studies did not find such differences [21].

The pancreas is anatomically divided into three segments: head, body, and tail. The pancreas head sits within a C-shape structure formed by the duodenum and joins with the pancreas body via the pancreas neck, a narrowing or ‘isthmus’ that bends around the superior mesenteric vessels. The pancreas neck is typically approximately 2 cm long and is commonly included as part of the head. The pancreas body spans from the left border of the superior mesenteric vein to the left border of the aorta, where it is joined to the tail. It is generally considered that the body-tail boundary is at the midpoint lengthwise of the two segments [22]. Other pancreas subsegment classification systems have been proposed for the purposes of surgical resection, based upon embryological foundations [22], [23]. Most studies of pancreas pathology using MRI have analyzed the images using regions of interest (ROIs), particularly a standard 3-ROI placement strategy targeting pancreatic head, body, and tail [20], [21], [24], [25], though some have placed an extra ROI in the pancreatic neck [19]. While ROIs have the advantage of avoiding artefactual regions, their choice of placement inevitably adds inter-observer variation that may obscure clinically important differences between pancreatic segments.

### 1.2. Pancreas subsegmentation

Pancreas segmentation, that aims to delineate the whole organ in 2-D or 3-D scans, has been proposed as an alternative analysis method to the 3-ROI placement strategy, which may improve observer-dependent bias and provide more advanced metrics for spatial assessment of chronic disease. However, such is the variability in size and shape of the pancreas that it is often considered too tedious to manually delineate in practice. The methods for pancreas segmentation proposed to date require widely differing amounts of user intervention. Manual segmentation is too costly and generally infeasible, especially in large databases such as the UK Biobank [26]. Metrics derived from pancreas segmentations are clinically important, for instance total pancreatic volume or the irregularity of the pancreas contour in the context of diabetes [27], [28]. Pancreas segmentations may also be used for subsequent characterization of the pancreas in functional or structural quantitative imaging data acquired separately during the same imaging session.

Automated pancreas segmentation methods that have been proposed to date have been based on traditional multi-atlas methodology or, more recently, convolutional neural networks [29]– [31]. The latter especially have reported remarkable accuracy. However, while these may provide whole-organ measurements, they do not characterize disease regionally by pancreas subsegments. One automated method for pancreas subsegmentation was reported based on k-means clustering [32], that was applied to pancreas motion analysis under radiation therapy. However, this method is dependent on initial seed points and multiple images from multiple breathing phases, and was not validated for accuracy. For these reasons, the validation of a robust, automated approach for pancreas subsegmentation is desirable, with potential to bridge the gap between currently available technology and standard clinical assessment.

Starting from a segmentation of a whole-organ, landmark-based approaches have been proposed for subsegmentation into the organ’s constituent parts, for example the Couinaud segments in the case of the liver [33]. In one approach, “landmarks” have been used to define planes of separation between the liver segments. However, landmark localization is relatively sensitive to noise and overall image quality. Other methods have addressed organ subsegmentation as a single task, in which segmentation models create a multi-label prediction, each label corresponding to an individual subsegment. For example, atlas-based segmentation uses image registration to propagate labels from a probabilistic template (constructed offline) to a target dataset. Multi-atlas segmentation (MAS) or deep learning (DL) segmentation methods may also be used, however, they typically need annotated parts individually on training subjects, and generally require large amounts of data. Some DL methods have drawn inspiration from traditional atlas-based methodology [34].

### 1.3. Purpose

In this work, a fully automated method based on groupwise registration is presented to subsegment the pancreas into its anatomical parts. Automatic delineations are assessed in comparison with those of human experts. Intra- and inter-rater variation of human experts is determined, that sets a benchmark for assessment of the automated method. We show that we can automatically segment pancreas parts in MRI, starting from a whole-pancreas segmentation, further enabling work studying disease heterogeneity. We validate the method retrospectively on a subset from the UK Biobank.

The purpose of this study is three-fold:

1. Propose a robust annotation protocol for delineating pancreas subsegments.
2. Introduce and validate an automated method for pancreas subsegmentation.
3. Show initial application of the method in regional assessment of pancreatic disease.

## 2. Materials and Methods

In Section 2.1, the data that were used for template creation are described, together with pre-processing of the training and validation data. The groupwise registration-based parts segmentation method is described in Section 2.2. In Section 2.3, we describe the validation experiment using expert annotations as reference. Section 2.4 shows the application of the method to a type 2 diabetes cohort of UK Biobank.

As benchmark, the method based on k-means proposed by Fontana et al. [32] was implemented for a single image (single ‘breathing phase’), choosing the initial cluster centroids using the k-means++ algorithm, and is referred to as the *k-means method* throughout this paper.

### 2.1. MRI data

MRI data from the UK Biobank imaging substudy was used. UK Biobank received ethical approval from the North West Multi-centre Research Ethics Committee (MREC) and written informed consent was obtained for all subjects. 100 subjects were used for template creation, 44 females and 56 males. All were nominally healthy subjects aged 50 to 70 with a mean age of 55 years for females and 57 years for males. The mean Body Mass Index (BMI) was 25.5 kg/m^2 for females and 27.1 kg/m^2 for males. An additional 50 subjects were used for validation, 21 males and 29 females, with a mean age of 53 and 57 years and a BMI of 25.9 and 26.5 kg/m^2, respectively.

All subjects had been scanned with a 1.5 tesla Siemens Aera (Siemens Healthineers, Erlangen, Germany) using a 2-point Dixon protocol covering neck to knee, acquired using 6 overlapping slabs and uploaded as Data-Field 20201. Only datasets from the first imaging session of UK Biobank (Instance 2) were used. Slabs were stitched together, and the resulting neck-to-knee volume was cropped to the abdominal region, resulting in a subvolume that generally included slabs 2, 3 and 4 (more details are available in [35]). Slabs 2, 3 and 4 each had voxel size 2.23 × 2.23 × 4.5 mm^3^ and matrix size 224 × 174 × 44.

The whole pancreas was delineated manually on all 150 training and validation datasets. Figure 1 shows 3-D renderings of the whole-pancreas segmentations for all subjects in both the template creation dataset and the validation dataset. The volumes and the corresponding whole-organ segmentations were resampled to 2 mm isotropic resolution. We also minimally co-registered the subjects by translating them to align their centroids. The centroid of subject 1 was used as a reference. The pre-alignment provided a better starting point for the nonlinear registration algorithm, both for template creation and method inference. Currently, the software is only compatible with isotropic images, identical image size and in approximate alignment with each other.

**Figure 1.**
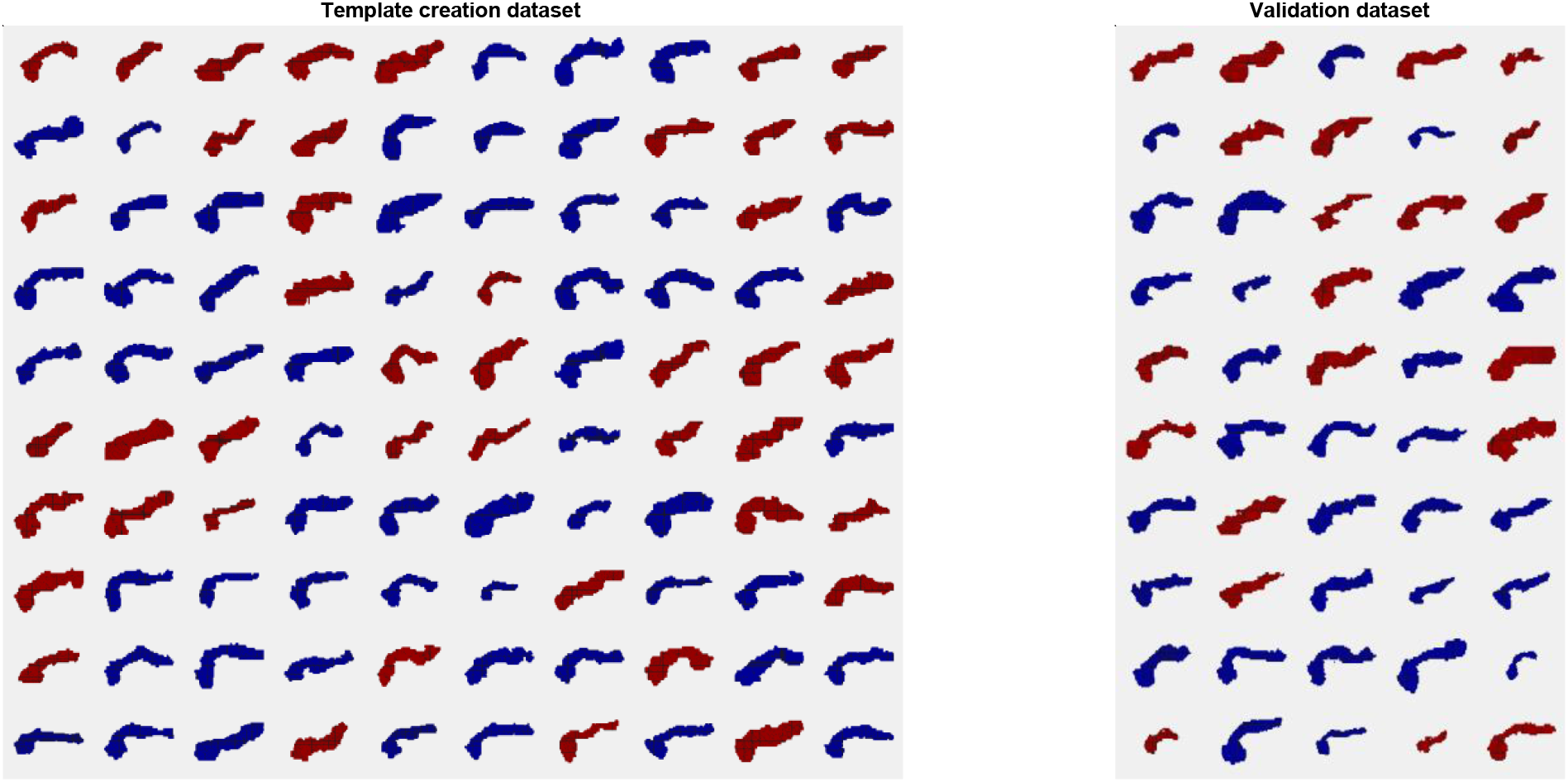
Whole-pancreas segmentations from the template construction (‘training’) dataset and the validation dataset, sorted by subjects’ age (females in red, males in blue).

### 2.2. Method description

An overview of the groupwise registration method is shown in Figure 2. The method takes a whole-pancreas segmentation as input, either delineated manually or with an automated approach. First, an average pancreas template is constructed offline using groupwise registration from the N=100 template creation dataset of whole-pancreas segmentations. Then, the pancreas parts (head, body, tail) are manually annotated on the constructed template, resulting in a pancreas parts template. Method inference (parts segmentation) is performed by registration of the pancreas parts template to a new target whole-pancreas segmentation. Then, the registered parts template labels are propagated to the target whole-pancreas segmentation, obtaining a pancreas parts segmentation for that subject. Offline parts template construction as well as parts segmentation inference steps are detailed in the following paragraphs.

**Figure 2.**
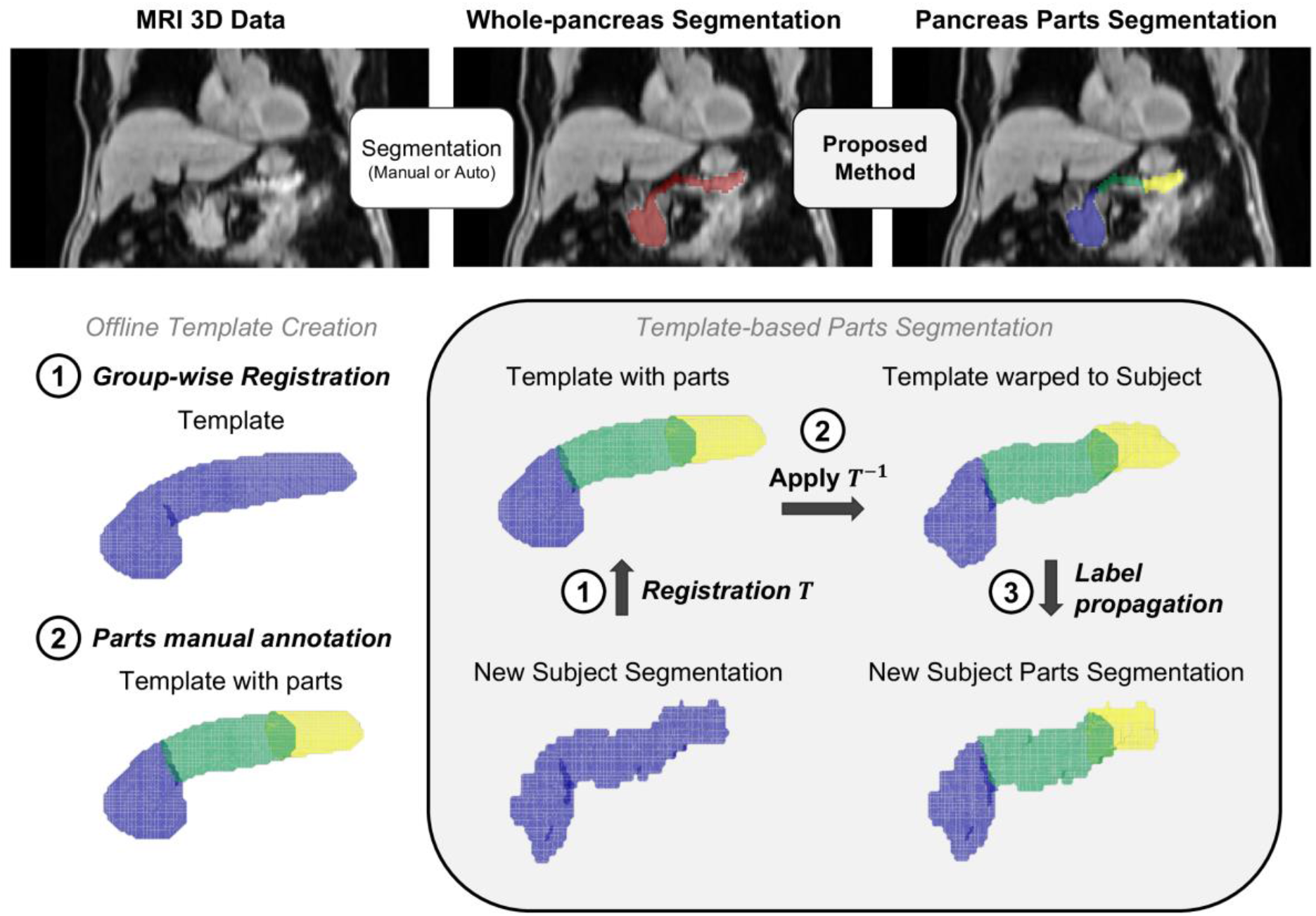
Method description. (top) Overall pipeline for whole-pancreas segmentation and parts segmentation. (bottom-left) (1) Offline groupwise registration of the whole-pancreas segmentation generated a population average (‘template’), on which (2) the parts were manually annotated (‘parts template’, head: blue, body: green, tail: yellow). (bottom-right) For a new subject, the method (1) computes a registration transformation from the subject’s segmentation to the template, (2) applies the inverse transformation on the parts template, and (3) propagates the warped parts template labels to the segmentation.

The backbone for template construction is the Large Deformation Diffeomorphic Metric Mapping (LDDMM) via Geodesic Shooting algorithm developed by [36] and available under the “Shoot” toolbox of the SPM12 software^1^. The toolbox uses diffeomorphic transformations to co-register all the template construction segmentations iteratively into a population average, i.e., the ‘template’ image. MATLAB R2021a (The MathWorks, Inc) and the batch processing capability of SPM12 were used to run template creation. A probabilistic template (0 to 1) was obtained from this step after 4 iterations, that was binarized by thresholding at 0.5.

Pancreas head, body, and tail were annotated on the template image. Note this template-based approach enables annotation of parts on the constructed template, instead of annotating each of the training subjects individually, thus requiring a single annotation step. The initial assumption was that this approach would not be significantly different from annotating each ‘training’ subject individually. One additional advantage of this annotation strategy is that some salient features appear on the template after groupwise registration, which correspond to the landmarks defining the pancreas subsegments. These landmarks may otherwise be difficult to identify in individual cases, and correct landmark identification is highly dependent on image quality. Annotation was performed by defining one boundary plane between head and body and another boundary plane between body and tail.

Given a whole-pancreas segmentation for a new subject, which can be either manually delineated or computed automatically, the method first computes a registration transformation from the subject’s whole-pancreas segmentation to the template (again initialized by aligning the centroid). The method then applies the inverse of that registration transformation onto the parts template. Finally, it propagates the labels of the warped parts template towards the whole-pancreas segmentation, obtaining a parts segmentation for that new subject.

### 2.3. Validation

A separate ‘validation’ dataset of N=50 subjects was gathered from UK Biobank with the same demographics as the template creation dataset. Automated pancreas subsegmentation was performed using the k-means method and the described groupwise registration method. Each subject’s whole-pancreas segmentation was also manually annotated into parts for reference, following a dedicated annotation protocol based on the 3-D ‘scalpel’ tool of ITK-SNAP^2^ [37]. The protocol instructed the drawing of 2 separation planes, one plane at the head-body boundary and one plane at the body-tail boundary, both as perpendicular to the pancreas centerline as possible. The protocol was distributed to 4 separate raters. One of the raters, who we refer to as *naïve observer*, had no prior experience with pancreas anatomy or pancreas imaging, so that the robustness of the annotation protocol to rater experience could be estimated. The three other raters, all of whom had expertise in medical imaging and familiarity with pancreas anatomy, were deemed “expert”. 10 of the 50 subjects were included at random twice in the dataset for the purpose of assessing intra-observer variability (referred to as annotation *a* and annotation *b*). This yielded a total of 60 annotations per rater. Inter-observer variability was also assessed by comparing annotations over multiple raters. The inter-observer performance may be used as a comparative benchmark for the automatic results.

Direct validation of automated pancreas subsegmentation was performed using generally accepted segmentation performance metrics, namely Dice Similarity Coefficient (DSC) and 95^th^ percentile Hausdorff Distance (95%HD), as well as the reported volume of each part. Intra-observer agreement, inter-observer agreement and ‘manual vs automated’ agreement were evaluated using Bland-Altman analysis [38] and Wilcoxon signed rank statistical testing. For the 10 subjects used in intra-rater variation assessment, a total of 30 datapoints (10 subjects × 3 expert raters) were generated and combined for the comparison: R1a vs R1b, R2a vs R2b, R3a vs R3b. For inter-observer variation assessment, 3 comparisons among raters were combined, R1 vs R2, R1 vs R3, R2 vs R3, with 50 parts segmentations in each comparison, yielding 150 datapoints. For manual vs automated (*Auto*), the following comparisons were performed and combined for each automated method separately: R1 vs Auto, R2 vs Auto, R3 vs Auto, each with 50 parts segmentations, that resulted in 150 datapoints. The volume of individual parts was also compared for its potential clinical relevance as a biomarker.

Indirect validation was also performed through quantification of pancreatic fat by parts, using the multi-echo gradient-recalled echo (GRE) 2-D single-slice data obtained from a separate breath-hold scan, uploaded as Data-Field 20260. The GRE scan has 2.5 × 2.5 × 6 mm^3^ resolution, 160 × 160 matrix size, 10 echoes, TE1 = ΔTE = 2.38 ms, TE10 = 23.8 ms, TR = 27 ms, and 20° flip angle. The median values from head, body and tail were reported after reslicing the parts segmentation onto the reconstructed proton density fat fraction (PDFF) map. A confounder-corrected magnitude-based chemical-shift encoding method [39] was used to reconstruct PDFF maps from the raw 10-echo GRE data. A spectral model from liver fat was used [40]. The 3-D parts segmentation volume was intersected with the 2-D PDFF map using the DICOM Reference Coordinate System information, as illustrated in Figure 3.

**Figure 3.**
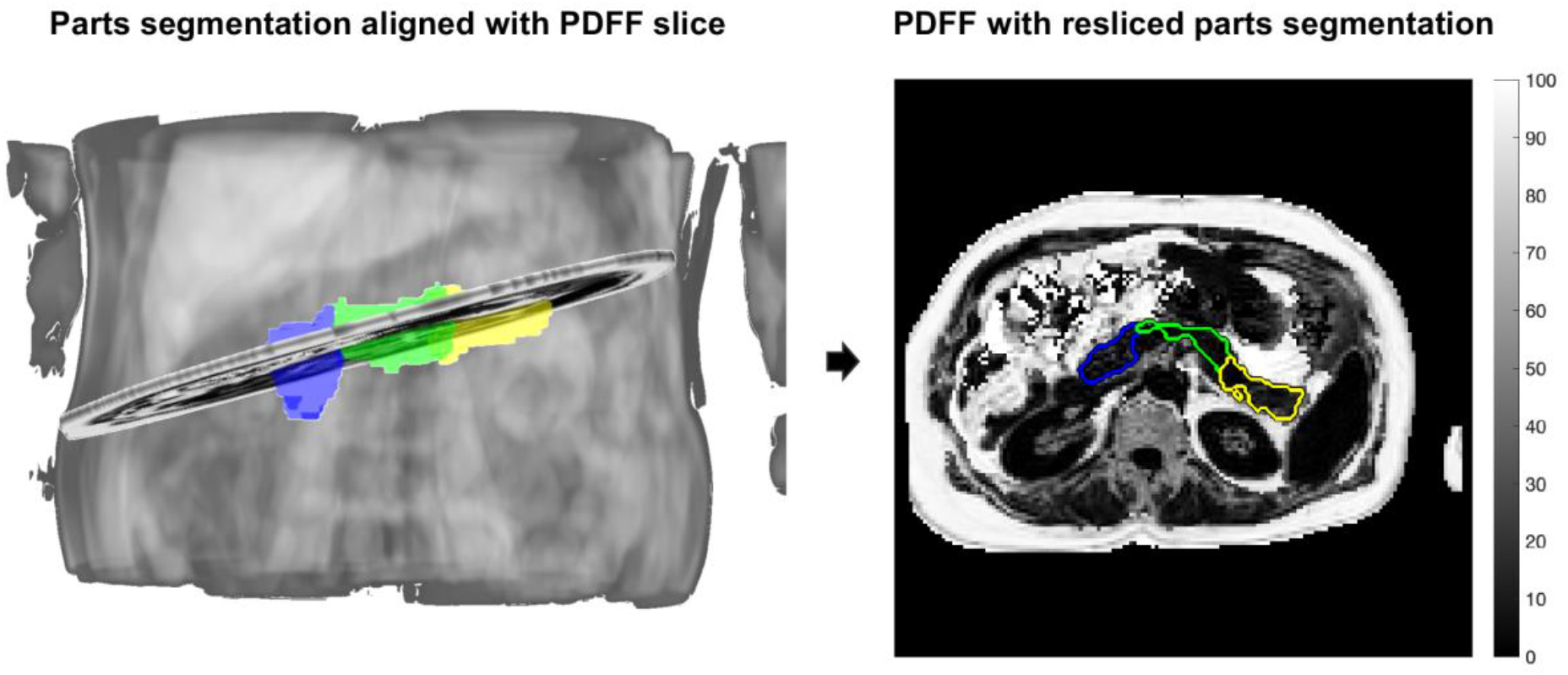
Pancreas segmentation (head: blue, body: green, tail: yellow) enables quantification of pancreas imaging biomarkers by parts, for example proton density fat fraction (PDFF), by intersection of the segmentation with the quantitative scan. PDFF, proton density fat fraction

Quantification differences were reported between the automated parts segmentation and the manual parts segmentations for each subject. Agreement in quantification was evaluated using Bland-Altman analysis. Segment masks with area of ≤30 pixels were excluded from the comparisons after a quality control (QC) step. The median PDFF of the segment masks was reported after excluding pixels with values exceeding 50%, followed by morphological opening with a disk structuring element of 3 pixels in diameter. The 50% PDFF threshold aimed to exclude non-parenchymal pancreatic tissue, for example surrounding visceral adipose tissue that could have been introduced due to slight subject motion between breath-holds.

### 2.4. Pancreatic fat quantification by parts in type 2 diabetes

As an initial exploration of fat heterogeneity in diseased subjects, a separate dataset of UK Biobank subjects was developed, comprising 390 triples of (1) self-reported type 2 diabetes mellitus (T2DM) subjects, (2) gender-, age- and BMI-matched non-diabetic subjects, and (3) gender- and age-matched non-diabetic subjects with chosen BMI of <25 kg/m^2. These groups of subjects will be referred to as: *T2DM, BMI-matched non-diabetics*, and *Matched low BMI non-diabetics* throughout this work. Age was matched to within 5 years, and BMI was matched within 1 point in all cases. A total of 390 × 3 = 1,170 subjects were collected.

Since manual whole-pancreas delineations were not available for these subjects, automated whole-pancreas segmentations obtained previously in [35] were used, predicted using the Attention U-Net model based on [31]. Some subjects had empty whole-pancreas segmentations that were excluded from the analysis. These were caused by failures in the model prediction, particularly in the presence of image artefacts. The groupwise registration-based automated parts segmentation method was run on the remaining whole-pancreas segmentations. The reslicing plus quality control approach explained in the previous section was run in order to measure median fat accumulation in the pancreatic head, body and tail. Pancreatic fat quantification by parts was compared between the 3 subject groups.

## 3. Results

### 3.1. Direct validation

Figure 4 illustrates manual and automated parts segmentations for the first 10 subjects in the validation set. 3-D renderings of manual parts segmentations from all 4 raters, as well as automated parts segmentations from the k-means method and the groupwise registration method are shown. The manual parts segmentations are similar across raters, including the naïve rater, with no major outliers. The manual parts segmentations look similar to the automated groupwise registration results, with no major outlier subjects, whereas the k-means method appears to overestimate the pancreatic head. These subjective judgements are quantified and confirmed below.

**Figure 4.**
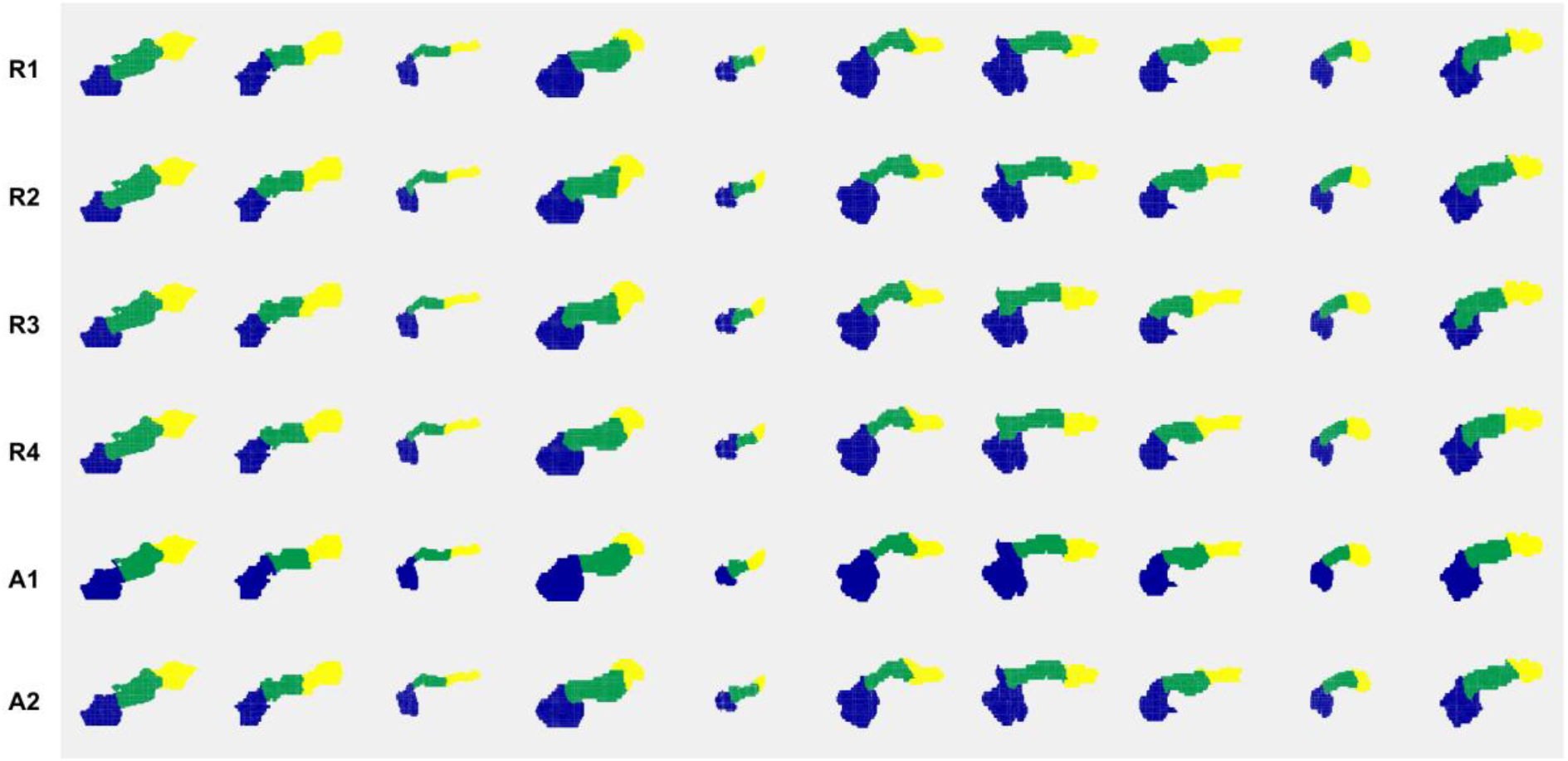
Qualitative evaluation of parts segmentations from Rater 1, Rater 2, Rater 3, Rater 4, Automated k-means method, Automated groupwise registration method, respectively (head: blue, body: green, tail: yellow). The first 10 subjects of the validation set are shown.

The robustness of the annotation protocol was tested against rater experience by comparing the segmentation performance in terms of DSC overlap of the naïve observer Rater (R) 4 vs themselves and vs the expert raters. The intra-rater agreement of a given expert observer R1 (R1a vs R1b) was not significantly higher than the intra-rater agreement of the naïve observer R4 (R4a vs R4b) (Wilcoxon signed rank test, right-tailed, Head: p=0.3848, Body: p=0.2158, Tail: p=0.3125). Similarly, the inter-observer agreement between 2 given expert observers, R1 vs R2, was not significantly higher than the inter-observer agreement between a given expert observer R1 vs the naïve observer R4 (Wilcoxon signed rank test, right-tailed, Head: p=0.9827, Body: p=0.9486, Tail: p=0.9352).

Table 1 reports intra-rater variation and inter-rater variation combined across the 3 expert raters and reported separately by pancreatic head, body and tail. Excellent intra-observer agreement (Dice overlap, Head: 0.982, Body: 0.940, Tail: 0.961, N=30) as well as inter-observer agreement (Dice overlap, Head: 0.968, Body: 0.905, Tail: 0.943, N=150) were observed in terms of segmentation performance. Intra-observer agreement was higher than inter-observer agreement.

**Table 1.** Direct validation metrics, expert raters combined. Dice Similarity Coefficient (DSC) and 95^th^ percentile Hausdorff Distance (95%HD in mm) are reported as mean ± standard deviation. Part volumes differences are reported in mL as bias [lower LoA, upper LoA]. Intra-observer agreement and inter-observer agreement are reported. LoA, limits of agreement

| *Experts combined* | **DSC** | **95%HD (mm)** | **ΔVolume (mL)** |
| --- | --- | --- | --- |
| **Intra-rater variation (n=10, N=30)** |  |  |  |
| head | 0.982 ± 0.014 | 0.609 ± 1.342 | 0.19 [-1.83, 2.21] |
| body | 0.940 ± 0.033 | 2.934 ± 2.420 | 0.08 [-3.95, 4.12] |
| tail | 0.961 ± 0.029 | 2.232 ± 2.509 | -0.28 [-4.25, 3.70] |
| **Inter-rater variation (n=50, N=150)** |  |  |  |
| head | 0.968 ± 0.022 | 2.171 ± 2.743 | 0.88 [-2.42, 4.19] |
| body | 0.905 ± 0.050 | 5.947 ± 4.033 | 0.06 [-5.93, 6.05] |
| tail | 0.943 ± 0.042 | 4.026 ± 4.121 | -0.95 [-5.63, 3.73] |

Table 2 reports ‘manual vs automated’ differences combined across the 3 expert raters for both the k-means method and the groupwise registration method. For the k-means method, significant differences were found between the combined inter-observer agreement and the combined ‘manual vs automated’ agreement, using DSC (Wilcoxon rank sum test, DSC, Head: p<0.001, Body: p<0.001, Tail: p=0.3965). For the groupwise registration method, no significant differences were found between the combined inter-observer agreement and the combined ‘manual vs automated’ agreement, using DSC (Wilcoxon rank sum test, DSC, Head: p=0.4358, Body: p=0.0992, Tail: p=0.1080). A statistically significant difference was found between the ‘manual vs auto’ agreement of the k-means method and the ‘manual vs auto’ agreement of the groupwise registration method for the head (Wilcoxon signed rank test, DSC, p<0.001) and body (p<0.001) segments, but not for the tail (p=0.6237).

**Table 2.** Direct validation metrics, expert raters combined. Dice Similarity Coefficient (DSC) and 95^th^ percentile Hausdorff Distance (95%HD in mm) are reported as mean ± standard deviation. Part volumes differences are reported in mL as bias [lower LoA, upper LoA]. Manual vs Automated agreement is reported for each of the existing automated methods. LoA, limits of agreement

| *Experts combined* | **K-means method** |  |  | **Groupwise registration method** |  |  |
| --- | --- | --- | --- | --- | --- | --- |
|  | **DSC** | **95%HD (mm)** | **ΔVolume (mL)** | **DSC** | **95%HD (mm)** | **ΔVolume (mL)** |
| **Manual vs Auto (n=50, N=150)** |  |  |  |  |  |  |
| head | 0.942 ± 0.045 | 6.196 ± 5.096 | -1.98 [-7.75, 3.79] | **0.965 ± 0.026** | **2.798 ± 3.755** | **-0.63 [-4.46, 3.20]** |
| body | 0.855 ± 0.120 | 8.899 ± 5.855 | 0.59 [-4.50, 5.69] | **0.893 ± 0.058** | **6.397 ± 3.890** | **0.08 [-5.69, 5.85]** |
| tail | **0.934 ± 0.054** | 4.870 ± 5.165 | 1.39 [-4.22, 7.00] | **0.934 ± 0.048** | **4.680 ± 4.130** | **0.55 [-4.81, 5.91]** |

### 3.2. Indirect validation

N=38 subjects had available the GRE sequence that enabled PDFF measurement out of the 50 subjects in the validation set. Note that, since the pancreatic PDFF scan is single-slice, the pancreatic head will not always present in the image due to variable slice positioning. Similarly, when slice position is too low, the pancreatic tail will not be visible. After processing and quality control, a total of 14 subjects with visible pancreatic head, 34 with visible body, and 29 with visible tail remained for quantification.

Figure 5 shows the Bland-Altman agreement in quantification for the inter-observer comparisons by pancreatic segments. Excellent agreement was observed between observers, for each part individually (head: bias=0.018, LoA=[-0.5, 0.5]; body: bias=-0.062, LoA=[-1.1, 1.0]; tail: bias=-0.019, LoA=[-1.6, 1.5]).

**Figure 5.**
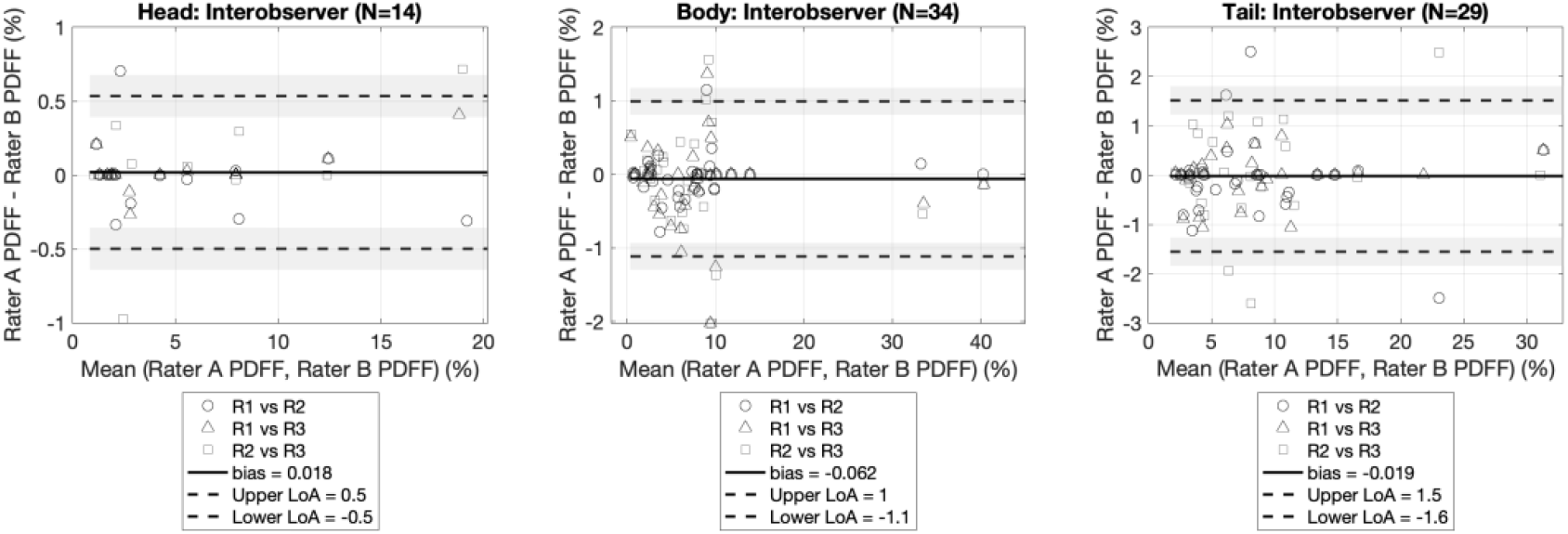
Inter-observer variation of derived PDFF quantification from the manual experts’ annotations.

Figure 6 shows differences in parts quantification between manual parts segmentations and the quantification derived from the automated parts segmentations, reported separately by parts, for both the k-means method and the groupwise registration method. For the groupwise registration method, the agreement between manual and automated segmentation quantification was excellent and comparable to the inter-observer comparisons. For the k-means method, differences were observed between the ‘manual vs auto’ agreement and the inter-observer agreement, especially in the head segment (bias=-0.409, LoA=[-2.5, 1.7]).

**Figure 6.**
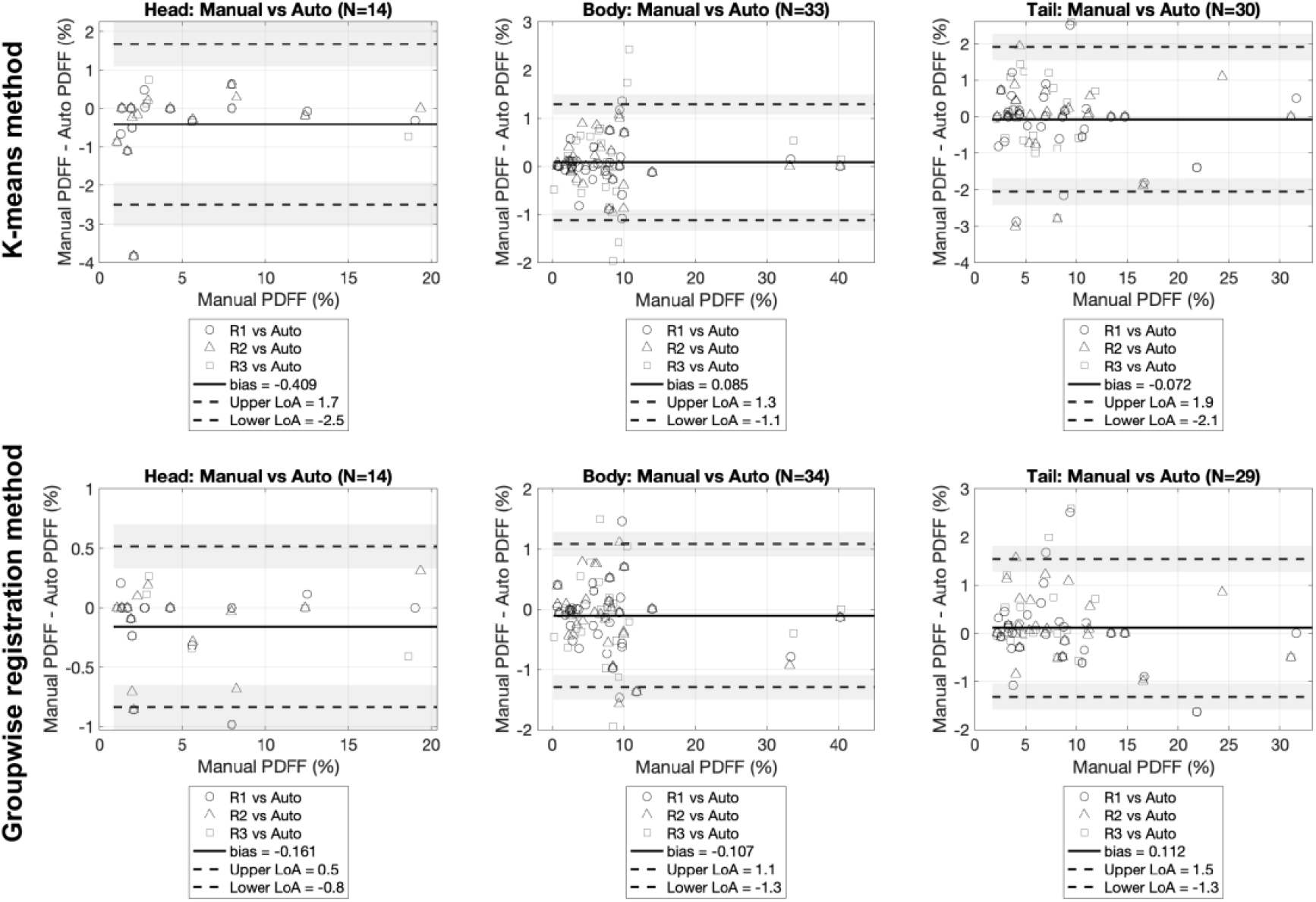
Manual experts’ annotations vs automated subsegmentations-derived PDFF quantification: differences by parts. ‘Manual vs auto’ comparisons are presented for both the k-means method and the groupwise registration method.

Figure 7 shows an example of a subject’s PDFF map with the resliced parts segmentations from Rater 1, Rater 2, Rater 3, the groupwise registration method and the k-means method. The k-means method appeared to overestimate the head segment, which may have caused the quantitative differences observed in the ‘manual vs auto’ comparison.

**Figure 7.**
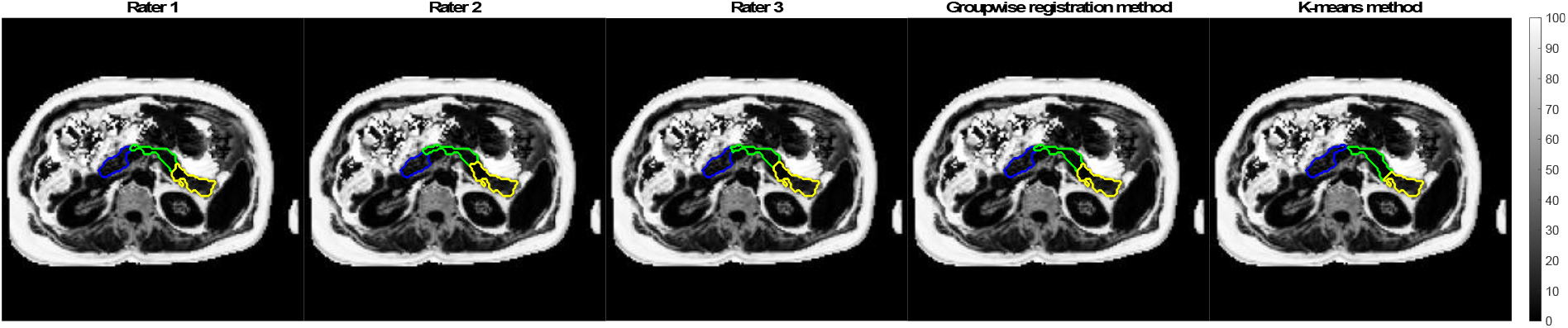
Example of regional pancreatic fat quantification for one subject in the validation set. Parts segmentation is shown on the PDFF image for Rater 1, Rater 2, Rater 3, Groupwise registration method, and K-means method, respectively (head: blue, body: green, tail: yellow). PDFF, proton density fat fraction

### 3.3. Pancreatic fat quantification by parts in type 2 diabetes

Figure 8 shows pancreatic fat quantification by pancreatic head, body, and tail for the 3 groups, (1) type 2 diabetics, (2) non-diabetics with matched age, gender and BMI, and (3) non-diabetics with matched age and gender and chosen low BMI. Total pancreatic fat is also included, which was obtained after combining all the part labels into a single ‘whole’ label.

**Figure 8.**
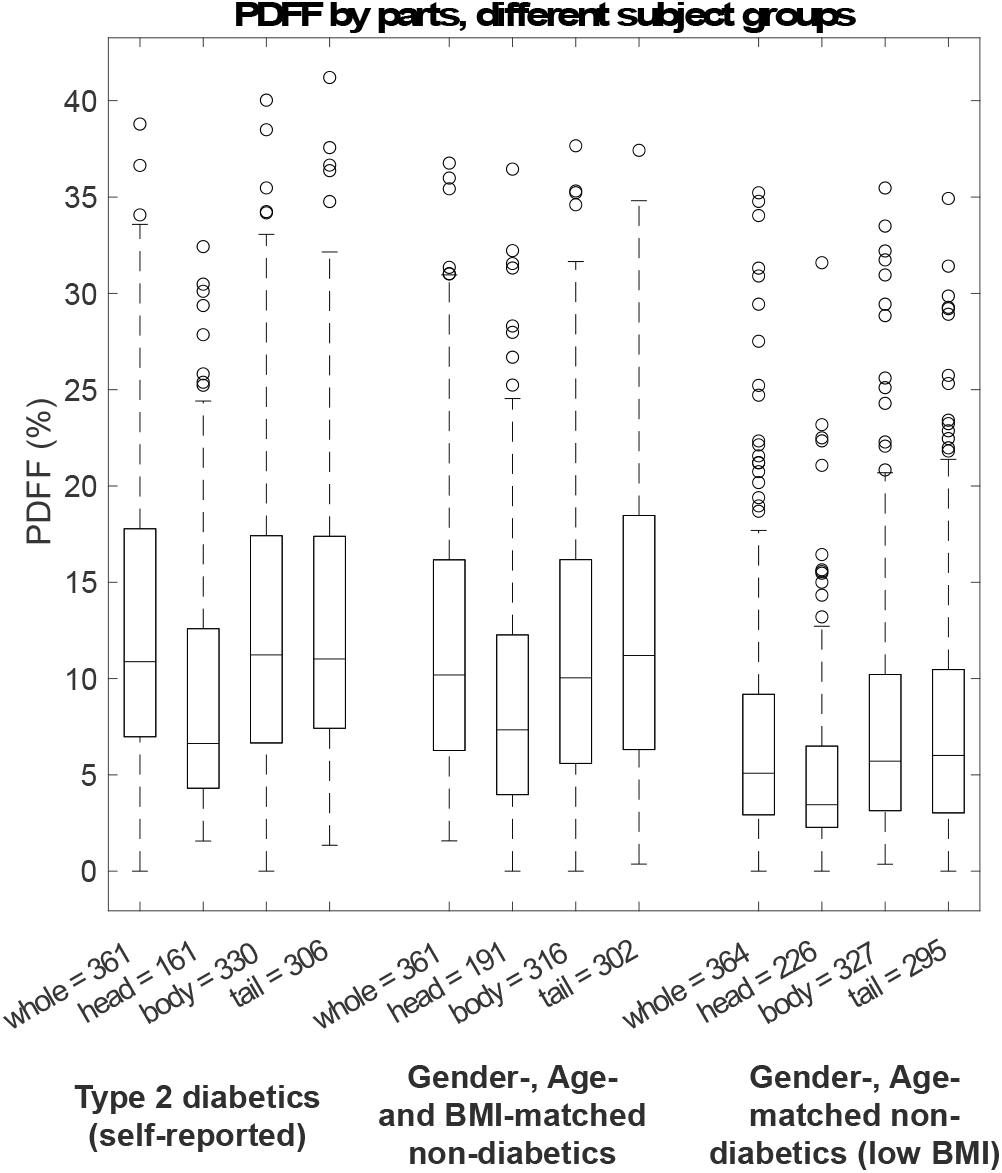
Pancreatic fat quantification by parts in groups from UK Biobank: (1) Type 2 diabetics (self-reported), (2) non-diabetics matched by gender, age, and BMI, (3) non-diabetics matched by gender and age and with low BMI. The number of segments available is displayed next to the part. BMI, body mass index

The Mann-Whitney U-test was used to compare quantification by parts across groups of subjects. The difference in PDFF of parts between T2DM and matched non-diabetic subjects with low BMI was significant when comparing all whole-pancreas PDFF, head PDFF, body PDFF, and tail PDFF (p<0.001 for all). The difference in PDFF of parts between T2DM and BMI-matched non-diabetics was only significant when comparing body PDFF (p<0.05), but not significant when comparing whole-pancreas PDFF (p=0.067), head PDFF (p=0.943) or tail PDFF (p=0.623).

Wilcoxon signed rank tests were used to evaluate differences in parts quantification between parts within each subject group. No significant difference between parts within groups was found, except for the BMI-matched cohort and the matched cohort with low BMI, where the head PDFF vs tail PDFF differences were significant (p<0.05 for both).

## 4. Discussion

This work presented and validated a fully automated method based on groupwise registration to subsegment the pancreas into its main anatomical parts: head, body, and tail. The method is based on a single population average or ‘template’ image and a single annotation stage on the template, which yields a parts template that may be used for pancreas subsegmentation in new subjects. The method was validated against manual annotations from expert observers in subjects from the UK Biobank imaging substudy and was compared to previously proposed unvalidated methodology based on k-means clustering [32]. Validation metrics included segmentation performance metrics as well as more clinically meaningful metrics like volume of parts and fat quantification by parts, which was obtained by intersecting the parts segmentations with proton density fat fraction (PDFF) maps. Then, as initial exploration of the clinical value of parts segmentation, the method was applied to a separate UK Biobank cohort including type 2 diabetics (self-reported) as well as gender-, age- and BMI-matched non-diabetic individuals, where spatial distribution of pancreatic PDFF was evaluated.

Note that automated whole-pancreas segmentation could have been used to generate both the template creation dataset and the validation dataset. However, by performing manual segmentations, any error introduced by whole-pancreas segmentation upstream of our method was minimized. This modular approach in which segmentation of the whole pancreas and the constituent parts are treated separately expedites validation of the subsegmentation method and allows for the introduction of improved whole-pancreas segmentation methods when they become available. In the final experiment, that showed the potential of parts segmentation, automated segmentations were used.

Excellent intra-rater as well as inter-rater agreement was observed among all raters for the proposed head, body, tail annotation protocol. This was true both for the 3 expert raters and the 1 ‘naïve’ rater, suggesting that the annotation protocol is robust, repeatable, and can be deployed by a range of raters.

Most literature quantifies imaging biomarkers by head, body, tail [20], [21], [24], as in the work presented here, though some researchers have considered the pancreatic neck separately in the quantification [19]. In early stages of developing the annotation protocol, four, rather than three, pancreas subsegments were considered by subdividing the head further into head and neck. This resulted in significant variation between raters. In any case, considering the image resolution of the PDFF map in UK Biobank, the pancreatic neck area would be comprised of few pixels, diminishing the reliability of neck PDFF quantification. Other acquisitions and applications may be more suitable for separate neck quantification, which we will revisit in future work. Other pancreas subsegmentation systems, for instance those incorporating embryological basis [22], [23], should also be considered in the future, for they may provide complementary regional assessment of the pancreas.

Excellent agreement was observed between the manual annotations and the automated groupwise registration method’s predictions, in terms of segmentation performance, subsegment volumes, and derived PDFF quantification. The agreement between expert raters and the automatic method suggests that the latter can be used in large datasets such as the UK Biobank. For this reason, the automated groupwise registration method was used in the subsequent experiment, which characterized regional quantification of fat in type 2 diabetics. One limitation of this is that template construction was performed using UK Biobank data comprising nominally healthy volunteers aged 50 to 70 with no self-reported diabetes of any type, though we plan to expand this cohort in future versions of the method. Applying the method to the type 2 diabetes cohort did not seem to impair method performance, based on a qualitative assessment (results not shown), though this needs careful evaluation.

As an initial exploration of the clinical application of our parts segmentation, we considered three matched groups: self-reported type 2 diabetes mellitus (T2DM) subjects, BMI-matched non-diabetics, and age- and gender-matched non-diabetics with low BMI. The significantly higher whole-pancreas PDFF in diabetics compared to non-diabetics has been reported previously [25]. However, we have shown that PDFF in the pancreatic body is significantly different between T2DM and BMI-matched non-diabetics, demonstrating the importance of parts segmentation beyond whole-pancreas measurements, which may obscure subtle but clinically important differences. One other study showed PDFF in the pancreatic tail to be most predictive for T2DM development within 4 years [20]. Our finding needs to be examined in more detail in future validation, for example using dedicated T2DM cohorts with longitudinal follow-up. The shown evidence of significant differences in pancreatic fat content between the pancreatic parts emphasizes the importance of segmentation-based approaches over ROI protocols, which should at least be ‘balanced’ when used, meaning they should target all pancreatic segments, for instance using multiple slices at different positions.

One advantage of groupwise registration methods is that they may be used for subsequent statistical analysis of biological variation across the population. Also, they often generalize robustly to various scan settings, compared to for instance deep learning methods, provided that the image resolution is in the same order. One criticism of templates is that they might average out differences between subjects. An approach that considers multiple templates based on major components of variation may be useful, for example clinical metadata information or imaging-based and radiomics features [29]. However, this increases the number of templates that need separate annotation. Evidently, in the extrema of this approach sit multi-atlas segmentation (MAS) methods, for which individual subjects in the training set need manual annotation of parts. Our approach seemed to balance well both performance and annotation efficiency. The template method’s predictions on the subjects it was trained with may provide good estimations of subsegmentations that could be used if labelling individual subjects is required, for example in MAS or deep learning methods, speeding the annotation process. The agreement observed between expert annotations and our automatic method supports this claim.

One method simplification could be introduced based on detecting the body-tail boundary using the pancreas segmentation centerline: the midpoint in length between the head-body boundary and the tip of the pancreatic tail would define the body-tail boundary more similarly to the anatomical definition used in this work, that is, “generally agreed to be located at the midpoint of the total length of the body and tail”, from [22]. We may also choose to fit each predicted boundary to a plane, similarly to the planes drawn in manual annotation, that is orthogonal to the pancreas centerline; in this scenario, the scalar distance between the manual boundary and predicted boundary planes may be used as the validation endpoint.

To date, we have studied regional differences for pancreatic PDFF, but note the method is suited to report differences in other biomarkers, such as T1, so long as the corresponding parametric maps are available within the imaging session.

## 4.1. Conclusion

This study demonstrated the feasibility of automated pancreas parts segmentation and downstream pancreatic imaging biomarker quantification by using groupwise registration of whole-organ segmentations to a template, and subsequent annotation of the template image. This enables segmental characterization of heterogeneous pancreatic disease.

## Data Availability

All data are available to approved researchers of UK Biobank.

## Acknowledgements

The authors thank Joel Robinson for help with delineations and feedback on the annotation protocol. We thank Perspectum Ltd for funding and computational resources. We also thank the Engineering and Physical Sciences Research Council (EPSRC) for the doctoral studentship award. This research was conducted using the UK Biobank Resource under Application Number 9914.

## Footnotes

1 https://www.fil.ion.ucl.ac.uk/spm/software/spm12/

2 http://www.itksnap.org/

